# Surveillance and correlation of SARS-CoV-2 viral RNA, antigen, virus isolation, and self-reported symptoms in a longitudinal study with daily sampling

**DOI:** 10.1101/2021.12.23.21268319

**Authors:** Gaston Bonenfant, Jessica Deyoe, Terianne Wong, Carlos G. Grijalva, Dan Cui, H. Keipp Talbot, Norman Hassell, Natasha Halasa, James Chappell, Natalie J. Thornburg, Melissa A. Rolfes, David Wentworth, Bin Zhou

## Abstract

The novel coronavirus pandemic incited unprecedented demand for assays that detect viral nucleic acids, viral proteins, and corresponding antibodies. The 320 molecular diagnostics in receipt of FDA emergency use authorization mainly focus on viral detection; however, no currently approved test can be used to infer infectiousness, i.e., the presence of replicable virus. As the number of tests conducted increased, persistent SARS-CoV-2 RNA positivity by RT-PCR in some individuals led to concerns over quarantine guidelines. To this end, we attempted to design an assay that reduces the frequency of positive test results from individuals who do not shed culturable virus. We describe multiplex quantitative RT-PCR (qRT-PCR) assays that detect genomic RNA (gRNA) and subgenomic RNA (sgRNA) species of SARS-CoV-2, including spike (S), nucleocapsid (N), membrane (M), envelope (E), and ORF8. The absolute copy number of each RNA target was determined in longitudinal specimens from a household transmission study. Calculated viral RNA levels over the 14-day follow up period were compared with antigen testing and self-reported symptoms to characterize the clinical and molecular dynamics of infection and infer predictive values of these qRT-PCR assays relative to culture isolation. When detection of sgS RNA was added to the CDC 2019-Novel Coronavirus Real-Time RT-PCR Diagnostic Panel, we found a qRT-PCR positive result was 98% predictive of a positive culture (negative predictive value was 94%). Our findings suggest sgRNA presence correlates with active infection, may help identify individuals shedding culturable virus, and that similar multiplex assays can be adapted to current and future variants.

## INTRODUCTION

The emergence and subsequent global transmission of severe acute respiratory syndrome coronavirus 2 (SARS-CoV-2), the causative agent of coronavirus disease 2019 (COVID-19), revealed current pitfalls in point of care diagnostics. Successful pandemic management depends on accurate and precise diagnostics with high throughput, fast turnarounds, and reproducibility. At the time of writing, one diagnostic has received full 510(k) clearance, while Emergency Use Authorization (EUA) from the U.S. Food and Drug Administration has been granted to 320 molecular (nucleic acid and antigen) and 90 serology diagnostics [1]. While active and previous infections can be documented using these authorized molecular and serology diagnostics, respectively, they are not designed to accurately assesses an individual’s infectivity, or ability to spread the virus.

Coronaviruses are enveloped, positive-sense RNA viruses responsible for up to one-third of annual common cold infections [2]. After cell entry, the viral RNA is released and the open reading frames (ORFs) ORF1a and ORF1ab are translated into the polyproteins pp1a and pp1ab, respectively, the latter resulting from a (−1) ribosomal frameshift [3]. Virus-encoded proteases process the two polyproteins into functional proteins responsible for viral RNA replication and immune evasion [4]. To produce the remaining structural and auxiliary proteins, the viral replicase complex utilizes the viral genome as a template to produce negative-sense RNAs for genome replication as well as shorter subgenomic RNAs (sgRNAs) through a process of discontinuous co-transcription [5, 6]. Each sgRNA contains the leader sequence from the 5’ end of the viral genome linked to a 3’ ORF encoding viral structural and nonstructural accessory proteins. Together, these sgRNAs form a set of 3’-nested RNA species. Because the process of sgRNA formation only occurs during genomic replication and transcription, sgRNA abundance has been proposed as a proxy of active viral replication and infectivity [7-9].

We describe here the development and characterization of quantitative real-time PCR (qRT-PCR) assays that detect SARS-CoV-2 genomic-specific RNA (gRNA) and sgRNAs for the spike, nucleocapsid, envelope, membrane, and ORF8 genes. Assays were validated with RNA extracted from supernatant of infected cells and synthetic RNAs for each analyte to quantify absolute copy numbers. We performed culture isolation with 452 longitudinal nasal specimens and used our multiplex assays, existing qualitative RT-PCR and antigen assays, and reported symptom data to characterize molecular kinetics of infection and infer our assays’ predictive value for viral culture isolation. Findings from this study suggest that research assays measuring sgRNA can be used to infer the presence of viable virus in specimens with qRT-PCR-detectable viral sgRNA and may inform public health recommendations regarding isolation of persons with SARS-CoV-2 qRT-PCR-positive results.

## MATERIALS & METHODS

### Specimen collection and initial processing

Between April-September 2020 (prior to emergence of B.1.1.7), households were recruited into a CDC-sponsored case-ascertained household SARS-CoV-2 transmission study in Nashville, TN, after identification of an infected index case [10]. Once enrolled, instructions demonstrating self-collection of anterior nasal specimens were provided to participants who also recorded symptom data daily for 14 consecutive days. Specimens were transferred to Vanderbilt University Medical Center (Nashville, TN) where they were processed and tested for SARS-CoV-2 using the CDC 2019-Novel Coronavirus Real-Time RT-PCR Diagnostic Panel (hereafter referred to as 2019-nCoV Assay; see Supplementary Information for additional details on specimen processing). Specimens passing the RNA quality assessment (Supplementary Information) were transported on dry ice to the Centers for Disease Control and Prevention (CDC) for further testing. The household transmission study protocol was approved by the Vanderbilt University Institutional Review Board. The CDC determined that the study was conducted consistent with applicable federal law and CDC policy (see 45 C.F.R. part 46; 21 C.F.R. part 56).

### Synthetic RNA design and qRT-PCR assay development

Plasmids, primers, and probes were designed using the SARS-CoV-2/Wuhan-Hu-1 sequence as a template (NCBI Reference Sequence: NC_045512.2). DNA fragments synthesized and cloned into pUC57 (GenScript) were designed with the T7 promoter upstream of the SARS-CoV-2 leader sequence, followed by the truncated ORF for a given transcript, and ended with an RNaseP gene fragment. Plasmid DNA linearized by restriction enzyme digestion was purified (Qiagen) and used as template for *in vitro* transcription (Promega P1320). RNA was purified by TRIzol-LS extraction and analyzed under denaturing conditions by agarose gel electrophoresis. Three singleplex assays per target were validated using synthetic RNA and RNA from supernatant of cells infected with SARS-CoV-2/WA1 (Supplemental Table 1; Supplemental Figure 1A & 2). The top performing primer set, determined by amplification efficiency, was selected for further validation and multiplex optimization (Table 1; Supplemental Figure 1B-E; Supplemental Table 3). Ct values for the two SARS-CoV-2 analytes and RNaseP in clinical specimens tested using the 2019-nCoV Assay correlated with gRNA and RNaseP Ct values using our multiplex assay (Supplemental Figure 2) [11, 12].

**Table 1:** qRT-PCR assays used for multiplexing and associated characteristics using synthetic RNA and viral RNA from cell culture supernatant

| Assay |  |  | Singleplex Assay |  |  | Multiplex Assay |  |  |
| --- | --- | --- | --- | --- | --- | --- | --- | --- |
|  |  |  | Slope | Calculated Efficiency (%) | R<sup>2</sup> Value | Slope | Calculated Efficiency (%) | R<sup>2</sup> Value |
| Synthetic RNA | MPlex1* | orf1ab_#1 | -3.314 | 100.3 | 0.9809 | -3.281 | 101.7 | 0.9971 |
|  |  | sg.Spike_#2 | -3.301 | 100.9 | 0.9885 | -3.354 | 98.7 | 0.9844 |
|  |  | sg.N_#1 | -3.319 | 100.1 | 0.9964 | -3.306 | 100.7 | 0.9920 |
|  | MPlex2 | sg.E_#4 | -3.232 | 103.9 | 0.9763 | -3.474 | 94.0 | 0.9934 |
|  |  | sg.M_#2 | -3.354 | 98.7 | 0.9773 | -3.574 | 90.5 | 0.9971 |
|  |  | sg.orf8_#2 | -3.466 | 94.3 | 0.9874 | -3.404 | 96.7 | 0.9972 |
| Virus from Infected Cells | MPlex1* | orf1ab_#1 | -3.301 | 100.9 | 0.9960 | -3.330 | 99.7 | 0.9943 |
|  |  | sg.Spike_#2 | -3.502 | 93.0 | 0.9568 | -3.413 | 96.3 | 0.9910 |
|  |  | sg.N_#1 | -3.264 | 102.5 | 0.9952 | -3.481 | 93.8 | 0.9771 |
|  | MPlex2 | sg.E_#4 | -3.214 | 104.7 | 0.9921 | -3.411 | 96.4 | 0.9298 |
|  |  | sg.M_#2 | -3.206 | 105.1 | 0.9538 | -3.522 | 92.3 | 0.9923 |
|  |  | sg.orf8_#2 | -3.292 | 101.3 | 0.9783 | -3.411 | 96.4 | 0.9703 |
|  | *This assay also included a primer and probe set for RNaseP |  |  |  |  |  |  |  |

**Figure 1.**
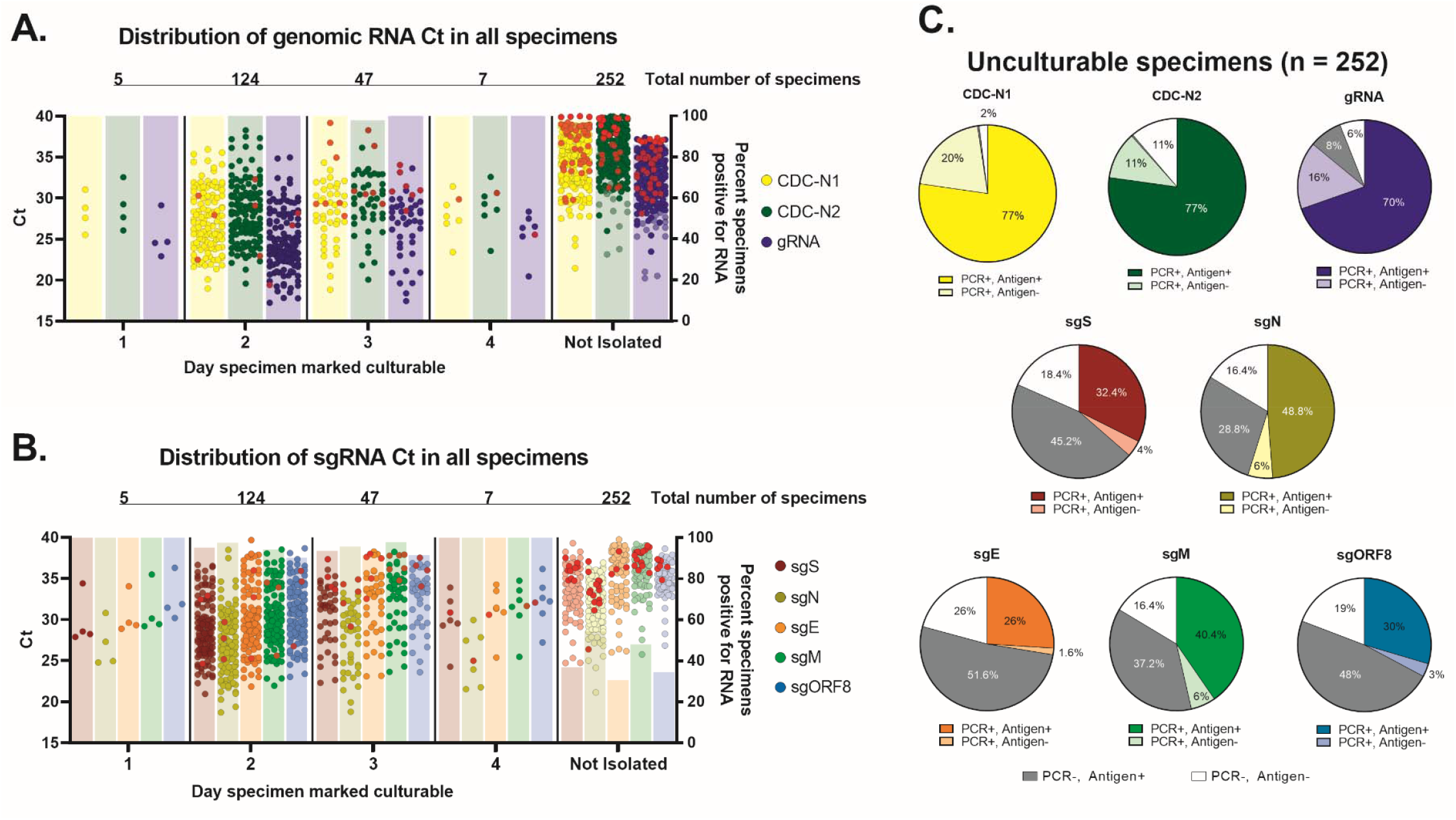
RNA and antigen results by culture isolation. A & B) The Ct values determined by qRT-PCR for all clinical specimens targeting genomic SARS-CoV-2 RNA (gRNA) and subgenomic RNA (sgRNA) are plotted by the indicated group along the x-axis. Above each dot plot are the total number of culturable or unculturable specimens isolated on a given day post inoculation. Bars plotted on the right y-axis indicate the percent of specimens with detectable RNA for each group. Solid red circles represent original clinical specimens negative for spike or nucleocapsid antigen by ELISA. C) For all unculturable specimens, antigen presence was compared to specimens testing positive or negative for the indicated analyte. Dark gray slices represent specimens that were qRT-PCR negative for the given analyte, but antigen positive for spike or nucleocapsid antigen, while white slices represent specimens qRT-PCR negative and antigen negative.

### Specimen processing, RNA detection, and antigen detection

Frozen respiratory specimens were thawed at room temperature and aliquoted into three pre-labeled tubes using aseptic technique: 100 µL for culture inoculation, 100 µL for antigen testing, and 100 µL in a third tube containing 400 µL AVL buffer (Qiagen). qRT-PCR was performed on a QuantStudio 6 Pro Real-Time PCR System (ABI). The aliquot for antigen testing was diluted 1:1 with specimen diluent and assayed for the presence of SARS-CoV-2 nucleocapsid or spike protein according to manufacturer’s recommendations (ABclonal RK04135, RK04136, and RK04159). Specimens were marked positive for antigen if the absorbance corresponded to a positive value per the standard curve equation. See Supplementary Information for further details on RNA extraction, qRT-PCR setup, and controls used for each.

### Inoculation of cells with clinical specimens and virus titration

Vero E6 cells (JCRB1819) stably overexpressing the transmembrane serine protease TMRPSS2 [31] were seeded into 24-well plates at a density of 1.8 × 10^5^ cells/well. On the day of infection, medium from plates was replaced with 200 µL infection medium. Skipping wells, 100 µL of undiluted clinical specimen was added dropwise to 12 wells and 100 µL infection medium to the remaining wells. One hour of virus adsorption with gentle agitation every 20 minutes was ended with the addition of 200 µL pre-warmed infection medium. Wells were visually inspected daily for five days; positive wells were designated as having >20% detached cells, at which time medium was collected. Wild-type SARS-CoV-2 from Washington state (SARS-CoV-2/WA1) was titrated by TCID50. Serial dilutions of virus prepared in virus diluent were transferred to a 96-well plate containing 80-98% confluent Vero E6 cells. Cytopathic effects were recorded between 3-7 days and TCID50 titer was calculated using Reed-Muench method [34]. The B.1.617.2 variant was titrated by plaque assay. Serial dilutions of virus prepared in virus diluent were transferred to 6-well plates containing 95% confluent Vero E6-TMPRSS2 cells. Plates were incubated for one hour with rocking every 10-15 minutes. Inoculum was removed and monolayers were overlaid with 1% cellulose solution. Assays were developed 48 hours later, and plaques were counted to calculate the titer (plaque forming units/mL) by dividing the average number of plaques for each dilution by the virus inoculum used per well. All incubation steps were performed at 37°C ± 5% CO_2_. See Supplementary Table 2 for frequently used reagent information.

### SARS-CoV-2 variant analyses and oligo consensus

Complete genomes absent of ambiguous nucleotides were randomly sampled from 01 July 2021 using the NCBI Nucleotide SARS-CoV-2 Data hub (https://www.ncbi.nlm.nih.gov/sars-cov-2/). Total sequences for B.1.1.7 and P.1 were bound to n = 2,000 to limit computational bandwidth. All available sequences in the database for B.1.351 (n = 351) and B.1.617.2 (n = 4,809) were included. Sequences were analyzed using Geneious “Test with Saved Primers”, an adaptation to Primer3 using the primers in Supplemental Table 2.

### Statistical analyses and data processing

Amplification efficiencies were calculated from the slope of the standard curve (*m*) using the following formula: efficiency (%) = [(10^−1/*m*^-1)x100]. Two-way ANOVAs were performed for statistically analyzing culture success in the presence and absence of given symptom(s) (Supplementary Figure 3C). Unpaired T-tests were performed for statistically analyzing culture success in the presence and absence of detectable antigen (Supplemental Table 4). The positive and negative predictive values of qRT-PCR relative to culture and vice versa were determined using the following equations:

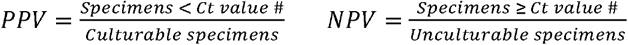

Consistency of qRT-PCR with culture referent:

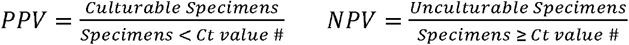

GraphPad Prism 8 was used to generate all figures.

## RESULTS

### Specimens isolated in culture and detection of viral analytes

We obtained 481 upper respiratory specimens self-collected between April-September 2020 from 79 people in 43 unique households. Based on initial RNA quality assessment (Supplementary Information), isolation of virus in cell culture was attempted with 452 specimens, of which 41% (187/452) were culturable (Cx^+^). Most Cx^+^ specimens were positive on day 2 (124/187, 66%) post-inoculation. All Cx^+^ specimens were positive for gRNA and most had detectable sgRNA. Among unculturable (Cx^-^) specimens (255/452, 56%), gRNA was detected in 88% (222/252) and sgRNA in 31-54% (77-137/252). Specimens with visible microbial contamination (n=10, 2%) or undetectable control RNaseP (n=4, 1.6%) were excluded from downstream analyses (Figure 1A & 1B).

Among Cx^+^ specimens, 92% (171/186) had detectable SARS-CoV-2 antigen (Ag^+^) (Figure 1). These specimens (n=171) had a mean Ct value ≤ 30 for each of the following unique targets: CDC-N1, CDC-N2, gRNA, sgS, and sgN. For Cx^+^ specimens lacking detectable antigen (Ag^-^; n=15), three had no detectable sgS or sgORF8, while gRNA and CDC-N1 were detected in all specimens. Significantly more gRNA and sgN was detected in Cx^+^/Ag^+^ specimens compared to Cx^+^/Ag^-^ (Supplementary Table 4). In Cx^-^/Ag^+^ specimens, sgRNA was detected in 34-63% (65-122/194), while CDC-N1 was detected in 100% (194/194). Of all analytes tested, only sgS, sgE, and sgORF8 had <50% qRT-PCR positive results in Cx^-^/Ag^+^ specimens (Figure 1C and Supplementary Table 4).

### Specimens cultured with high Ct and extended shedding of viable virus

Ten specimens with Ct values >34 for CDC-N1 were Cx^+^ (Ct^high^/Cx^+^), while seven specimens with Ct values <30 for CDC-N1 were Cx^-^ (Ct^low^/Cx^-^) (Supplementary Table 5). At the time of collection, a similar percent of participants from both groups were symptomatic (7/10, 70% for Ct^high^/Cx^+^; 5/7, 71% for Ct^low^/Cx^-^). Most participants reached peak viral concentrations in their specimens within the testing period (Figure 2A-C). Four individuals (4/58, 6.9%) produced Cx^+^ specimens for more than seven days. Symptoms were reported by these four participants at the time of collection for 73% (8/11) of specimens, of which all were Ag^+^ and all sgRNA analytes were detected in each specimen except for sgE in one. Symptom reporting and frequency were similar between individuals with less than or greater than six days of culturable virus (Supplementary Table 6).

**Figure 2.**
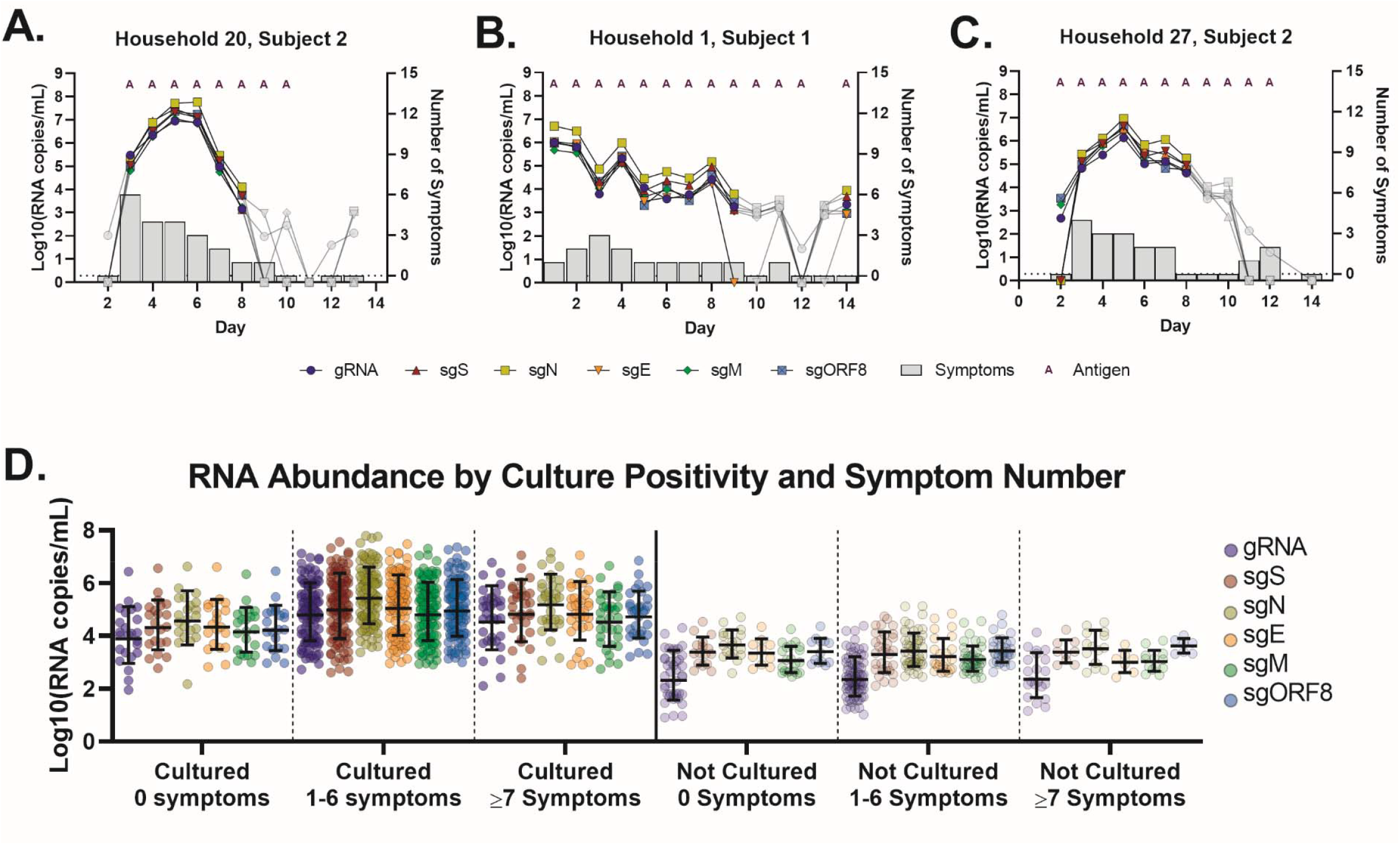
RNA fluctuation during the molecular time course of infection. A, B, & C) Calculated RNA copies/mL are plotted by day of sampling along the x-axes for selected participants. Gray data points indicate unculturable specimens, while colored symbols indicate the culturable specimens. The number of self-reported symptoms shown in gray bars are plotted on the right y-axes. Specimens with a positive test for antigen are represented with an “A” at the top of each data point, while the absence of “A” indicates a negative test for antigen. Specimens with undetectable RNA by qRT-PCR for any given analyte were arbitrarily assigned a value 0. D) Specimens were grouped by number of symptoms reported and culture result. Standard curves generated using synthetic RNA for each analyte were used to determine the number of RNA molecules per reaction. Specimens with no detectable RNA are not plotted and did not factor into the mean or 95% confidence interval.

### Detection of sgRNA increases probability that specimens contain viable virus

We compared Ct values of Cx^+^ specimens (referred to as true positives) with Ct values of Cx^-^ specimens (referred to as false positives) to assess the predictive value of our assay. RT-PCR data from the 2019-nCoV Assay initially performed on specimens was used as the comparator for our multiplex assays. An RT-PCR positive result from the 2019-nCoV Assay was 50.4% (181/359) predictive of a Cx^+^ specimen, while a RT-PCR negative result was 99% (68/69) predictive of an Cx^-^ specimen (Table 2). We examined stepwise increases in Ct values for unique RNA targets from our multiplex assays and evaluated the consistency of culture to qRT-PCR data (Supplemental Figure 4A). When using unique Ct cutoffs for gRNA, sgS, and sgN, we achieved a PPV of 96% (159/166) and a NPV of 91% (239/262). We found a Ct cutoff of 35 for both 2019-nCoV Assay analytes with a Ct cutoff of 36 for sgS led to a PPV of 98% (166/170) and a NPV of 94% (242/258) (Table 2; Supplemental Figure 4B).

**Table 2:** Correlation of qRT-PCR data with specimen isolation

| Cutoff | Consistency of qRT-PCR to Culture |  | Consistency of Culture to qRT-PCR |  | Overall Predictive Value* |
| --- | --- | --- | --- | --- | --- |
|  | PPV<br>qRT-PCR<sup>+</sup>/Cx<sup>+</sup> | NPV<br>qRT-PCR<sup>-</sup>/Cx<sup>-</sup> | PPV<br>Cx<sup>+</sup>/qRT-PCR<sup>+</sup> | NPV<br>Cx<sup>-</sup>/qRT-PCR<sup>-</sup> |  |
| N1 < 40<br>N2 < 40 | 181/182 (99.5%) | 68/246 (27.6%) | 181/359 (50.4%) | 68/69 (98.6%) | 74.5% |
| sgS < 31 | 112/182 (61.5%) | 246/246 (100%) | 112/112 (100%) | 246/316 (77.8%) | 89.9% |
| gRNA < 35 | 182/182 (100%) | 118/246 (48.0%) | 182/310 (58.7%) | 118/118 (100%) | 79.4% |
| gRNA < 30<br>sgS < 35<br>sgN < 34 | 159/182 (87.4%) | 239/246 (97.2%) | 159/166 (95.8%) | 239/262 (91.2%) | 93.5% |
| N1 < 35<br>N2 < 35<br>sgS < 36 | 166/182 (91.2%) | 242/246 (98.4%) | 166/170 (97.6%) | 242/258 (93.8%) | 95.7% |
| PPV: Positive predictive value<br>NPV: Negative predictive value<br>*Overall predictive value = (PPV + NPV from consistency of culture to qRT-PCR)/2 |  |  |  |  |  |

### Self-reported symptom progression and temporal decline in viral RNA

Most Cx^+^ (152/184, 83%) and Cx^-^ (150/236, 64%) specimens were from participants reporting one or more symptoms (Figure 2D). The three most commonly reported symptoms from participants with Cx^+^ specimens were fatigue (106/183, 58%), headache (56/101, 56%), and nasal congestion (86/183, 47%), while the top three from participants with Cx^-^ specimens were anosmia (87/207, 42%), headache (50/119, 42%), and fatigue (96/236, 40%). Data from each participant were organized longitudinally to analyze RNA levels and symptom data over the time course of infection (Figure 3A). Viral RNA levels and the number of participants reporting nasal congestion, fatigue, runny nose, and shortness of breath peaked between the 2^nd^ and 5^th^ day. As RNA levels decreased, the proportion of participants reporting symptoms followed (Figure 3B; Supplemental Figure 3A). Regardless of symptom presence, RNA abundance was significantly higher in all Cx^+^ specimens compared to Cx^-^ specimens. In general, RNA abundance was greater in symptomatic individuals than asymptomatic individuals within the same group. Interestingly, we found significantly less gRNA and sgM in Cx^-^ specimens that originated from participants self-reporting fever with fatigue or fever with headache (Supplemental Figure 3).

**Figure 3.**
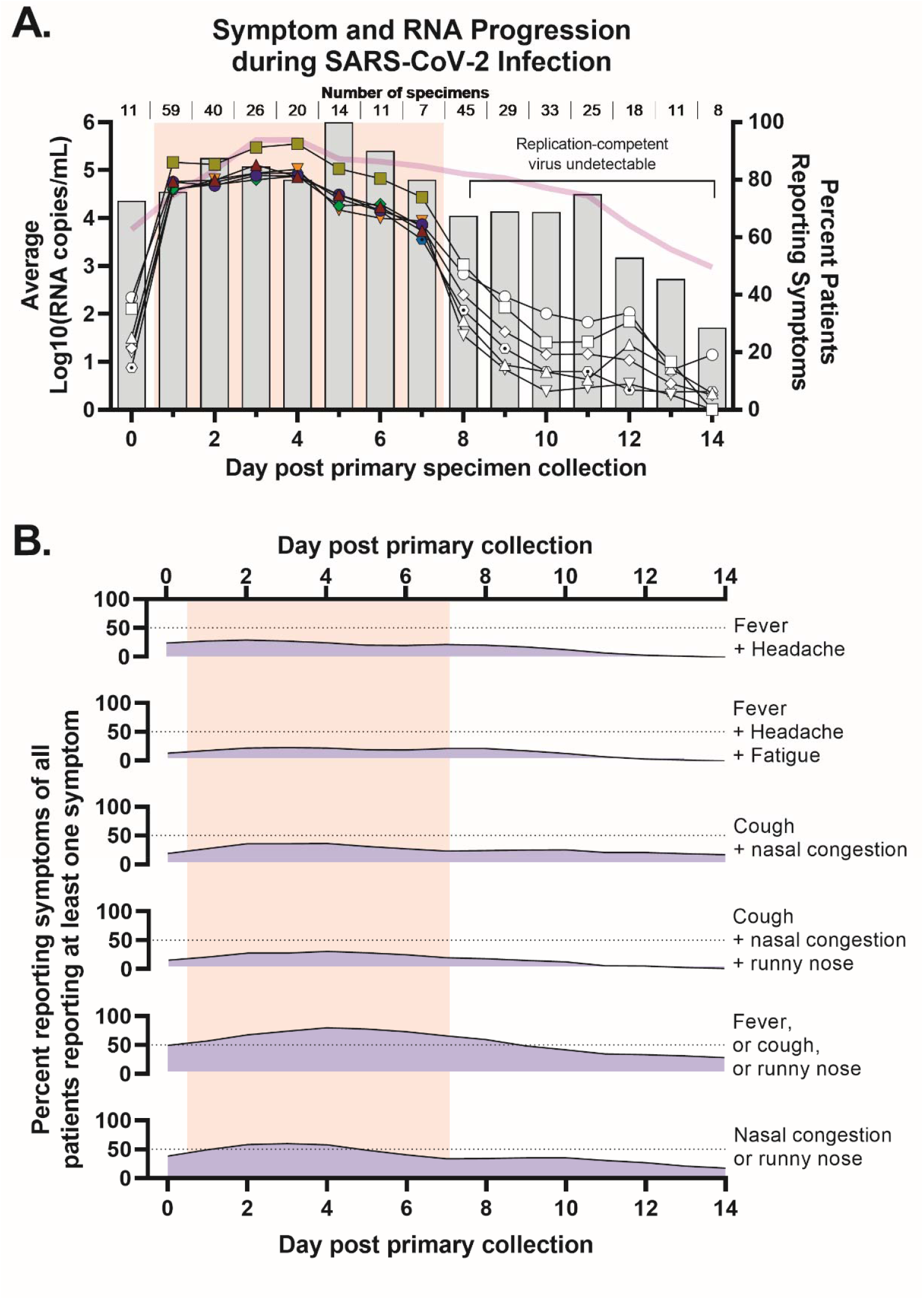
SARS-CoV-2 RNA abundance and symptoms follow similar trend. A) The average RNA copies/mL on each day of disease progression are plotted on the left y-axis and percent of patients reporting symptoms on a given day on the right y-axis. The number of patient specimens for each day graphed are listed under the title. Colored symbols and transparent red box represent culturable specimens and white symbols represent unculturable specimens. The semi-transparent purple line represents the average percent of antigen positive specimens for each day. B) X-axis follows same designation as Figure 3A. The percent of patients reporting a combination of symptoms out of only those who reported one or more symptoms are graphed. Light gray dotted line indicates 50%. Solid black lines are 2^nd^ order smoothing (4 neighbors). Transparent red box indicates days of self-collection patient specimens rendered culturable virus. Data points with less than 6 patient samples (e.g., patient specimens with consecutive culture positive results for 8-14 days or specimens culture negative more than two days prior to the first culture positive specimen for the same patient) were not included in this figure or data analyses.

### Application to B.1.617.2 variant

Sequence analysis on a subset of SARS-CoV-2 specimens showed less than 1.4% mismatches in the binding region for all gRNA oligos and sgN reverse primer and probe sites. While a similar percentage of sequences contained mismatches for the sgS reverse primer site in B.1.1.7, 32% of B.1.351 sequences, 99% of P.1 sequences, and 99.6% of B.1.617.2 sequences contained 1-2 mismatches at the 3’ end of the reverse oligo (Supplementary Figure 5A). Using B.1.617.2 sgS-specific oligos, we observed similar amplification efficiencies and sensitivity when using synthetic RNA encoding the SARS-CoV-2/WA-1 spike sequence (data not shown) and total RNA extracted from supernatant of SARS-CoV-2/WA-1- or B.1.617.2-infected cells (Supplementary Figure 5B, 5C). Both multiplex mastermixes performed similarly with RNA extracted from 50 clinical specimens originating from a recent infection cluster (Supplementary Figure 5D). Compared to the original batch of specimens collected in mid-2020, most of the recent specimens were culture-positive one day post infection (Supplemental Figure 5E).

## DISCUSSION

In this study, we evaluate two multiplex qRT-PCR assays and provide unique insight into the association with and variability between the characteristics of SARS-CoV-2 infection. After validating our testing method, we used longitudinal specimens collected from individuals soon after their infection to document a temporal relationship between symptom presence, infectious virus, RNA abundance, and antigen presence. We found SARS-CoV-2 sgRNA abundance more strongly correlated with specimens containing culturable virus compared with positive antigen tests. On average, the abundance of each viral RNA peaked on the third and fourth day of infection, while the percent of participants reporting symptoms peaked on the fifth. The number of self-reported symptoms from participants with Cx^+^ specimens was often greater than those among Cx^-^ specimens, though we found no significant difference in the number or type of symptom reported between individuals with less than or greater than 6 days of culturable virus.

The recommendation made early during the COVID-19 pandemic by the World Health Organization (WHO) that previously infected individuals end quarantine after two negative RT-PCR tests [13] led to prolonged isolation for individuals who persistently tested positive after symptoms subsided and those who tested positive after receiving two negative tests [14]. In this report, 41% (187/452) of specimens we assayed were Cx^+^ even though most (419/452, 93%) were positive by RT-PCR using the 2019-nCoV Assay. While testing positive for SARS-CoV-2 by RT-PCR indicates infection, a significant proportion of RT-PCR positive persons, encouraged to self-isolate based strictly on Ct values, may not be infectious. Diagnostic sgRNA testing and the value it may afford in limiting isolation or confirming persistently positive cases has been studied [15-18] and evidence exists supporting sgRNA abundance correlates with infectivity [9, 19-22]. All five sgRNA analytes from our multiplex qRT-PCR assays were detected in 90% (167/186) of Cx^+^ specimens compared to 19% (49/252) of Cx^-^ specimens. Our assay reduced the frequency with which unculturable specimens containing detectable viral RNA were labeled positive and maintained a similar percent of false negatives (Cx^+^/Ct^-^) as the 2019-nCoV Assay, suggesting sgRNA presence correlates with active infection and may help identify individuals shedding culturable virus. Of course, these conclusions may have some limitations based on the following assumptions: 1) virus isolation in culture directly correlates to a person’s infectivity; 2) viral RNA abundance correlates with infectious units; 3) all sampling and storage methods lead to similar quantities of specimen available for testing; 4) the relationship between viral RNA abundance and infectivity is consistent and robust to factors negatively influencing RNA or virion integrity. While one or more of the foregoing might not hold true, sgRNA is nonetheless produced during viral replication and would likely indicate active infection.

Detection of sgRNAs can vary dramatically depending on specimen and tissue source [20], storage conditions [30], symptom presence and severity at the time of sampling [32], patient demographics, and overall medical history [33]. In patient specimens, sgRNA can be detected for more than two weeks after initial detection and has been found for up to 162 days by PCR [20, 23]. Some specimens we tested were Cx^-^, yet positive for sgRNA. One proposed mechanism ascribes nuclease-resistance and structural stability as the basis of sgRNA [24]. Replication of SARS-CoV-2 and sgRNA production likely occur in or on double-membrane vesicles [25], which when associated with sgRNA can promote nuclease-resistance. Alternatively, extended, or recurrent sgRNA detection has been reported in immunocompromised individuals and therefore can be a sign of a persistent, active infection [26-27]. Thus, any diagnostic interpretation related to sgRNA presence must be considered from a broad perspective that includes symptom features, co-morbidities, and other factors influencing infection outcomes.

Viral antigen tests that utilize lateral flow mechanisms have been widely deployed for surveillance. In addition to rapid time-to-result, scalability, and low cost, results from these assays correlate with culture data [28]. We found a strong correlation between Ag^+^ results by ELISA and successful viral culture, but observed a high frequency of Cx^-^/Ag^+^ specimens. A recent assessment of five SARS-CoV-2 antigen assays identified a false positive rate between 1-39% compared to RT-PCR and culture isolation but relatively few false negatives compared to RT-PCR [29]. A growing concern has centered on SARS-CoV-2 genetic variants and testing outcomes. Because assays described in this article were developed prior to emergence of these variants, we evaluated test performance using 50 clinical specimens collected when the Delta variant accounted for more than 99% of sequenced genomes and found the original and adjusted multiplex assay led to similar Ct values. Although sensitivity and specificity toward the B.1.617.2 variant using the original multiplex assay was comparable to our data with the prototype virus, we cannot exclude the possibility that other circulating or future variants will perform as well.

In studies described here, we characterized two multiplex qRT-PCR assays for the detection of SARS-CoV-2 gRNA and sgRNAs. We showed that sgRNA abundance more strongly correlates with isolation of virus from specimens in culture than the presence of viral antigen. Over time, the proportion of participants reporting fever, cough, or runny nose correlated with viral RNA abundance. Although RNA levels in some Cx^-^ specimens approximated loads in Cx^+^ specimens, most Cx^-^ units harbored at least one log less of each analyte RNA. Moreover, the abundance of sgN was on average greater than gRNA abundance in Cx^+^ vs. Cx^-^ specimens. When applied to a real-world setting, symptom presence combined with detection of at least one sgRNA target with or without gRNA may help accurately identify individuals who are infectious, thereby guiding appropriate isolation to limit transmission while mitigating the personal, social, and economic impact of unnecessary isolation.

## Data Availability

All data produced in the present study are available upon reasonable request to the authors

## ACKNOWLEDGEMENTS

The authors extend their gratitude to participants who willingly self-sampled for the duration of this study, CDC staff and personnel for their continued support during the COVID-19 pandemic, and the physicians and other public health professionals responsible for educating, testing, treating, and tracing infected persons.

## FIGURE LEGENDS

**Supplemental Figure 1:**
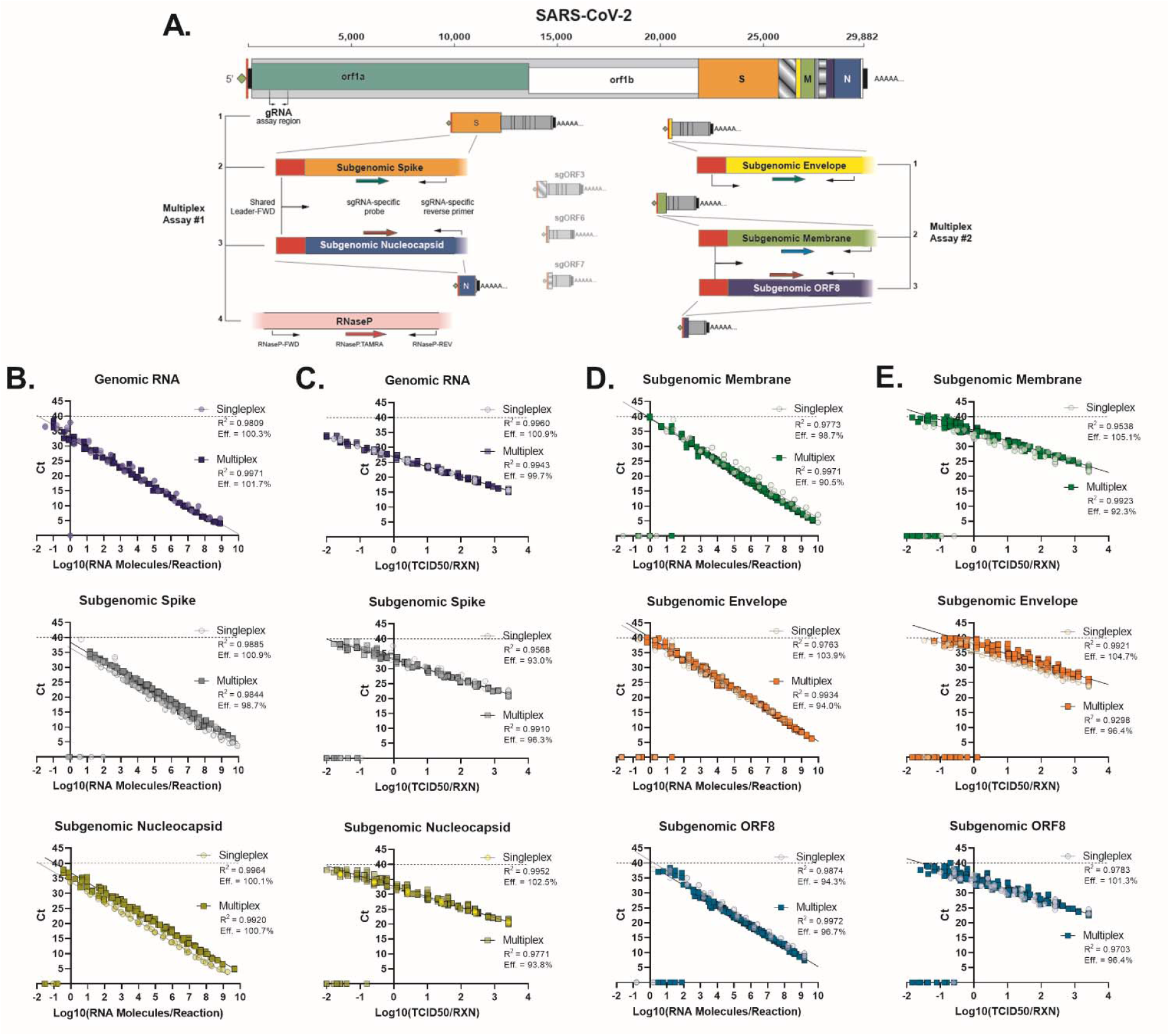
SARS-CoV-2 genome architecture and multiplex assay design with validation using synthetic RNA and virus from infected cells. A) Schematic illustrating the full-length genome with subgenomic RNA species used (saturated colors) in multiplex assays and those not used (faded colors). B & D) Ct values plotted against the synthetic RNA for a given analyte. C & E) Ct values plotted against the approximate tissue culture infectious dose for the indicated analyte using total RNA from supernatant of VeroE6 cells infected with SARS-CoV-2. Listed underneath the assay type are the R^2^ and primer efficiencies as determined by the slope for the linear trendline.

**Supplemental Figure 2:**
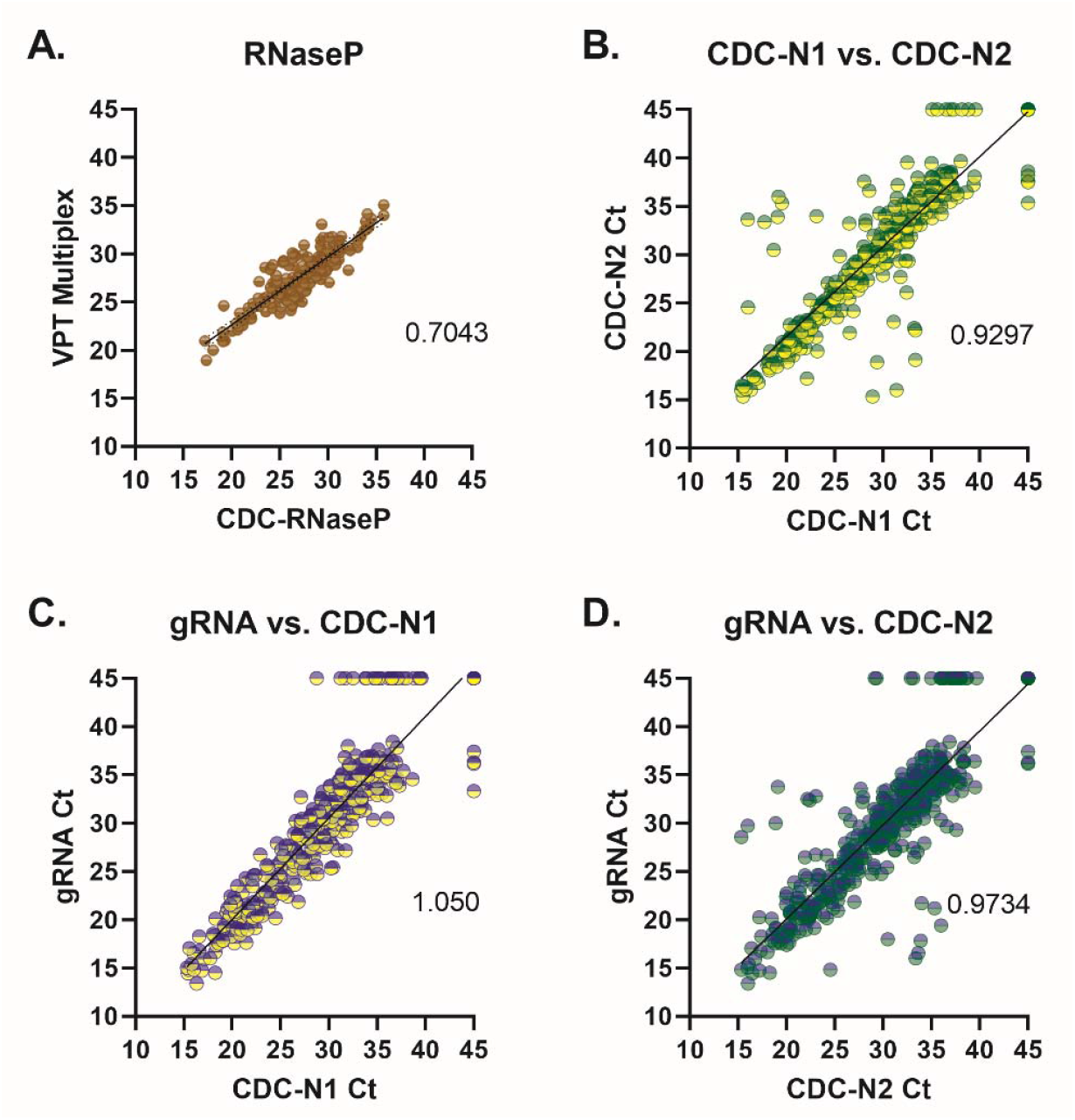
Validation of multiplex assay using previously tested clinical isolates. A) RNaseP Ct values for controls plotted on X-axis and RNaseP Ct value from multiplex assay on Y-axis. B) Inherent variation in N1 and N2 assays. C & D) gRNA from multiplex plotted against N1 and N2. The slope of the best-fit line is inset in each graph. All samples were subjected to freeze-thaw at least once and multiplex assays were performed on a different instrument from control assays.

**Supplemental Figure 3.**
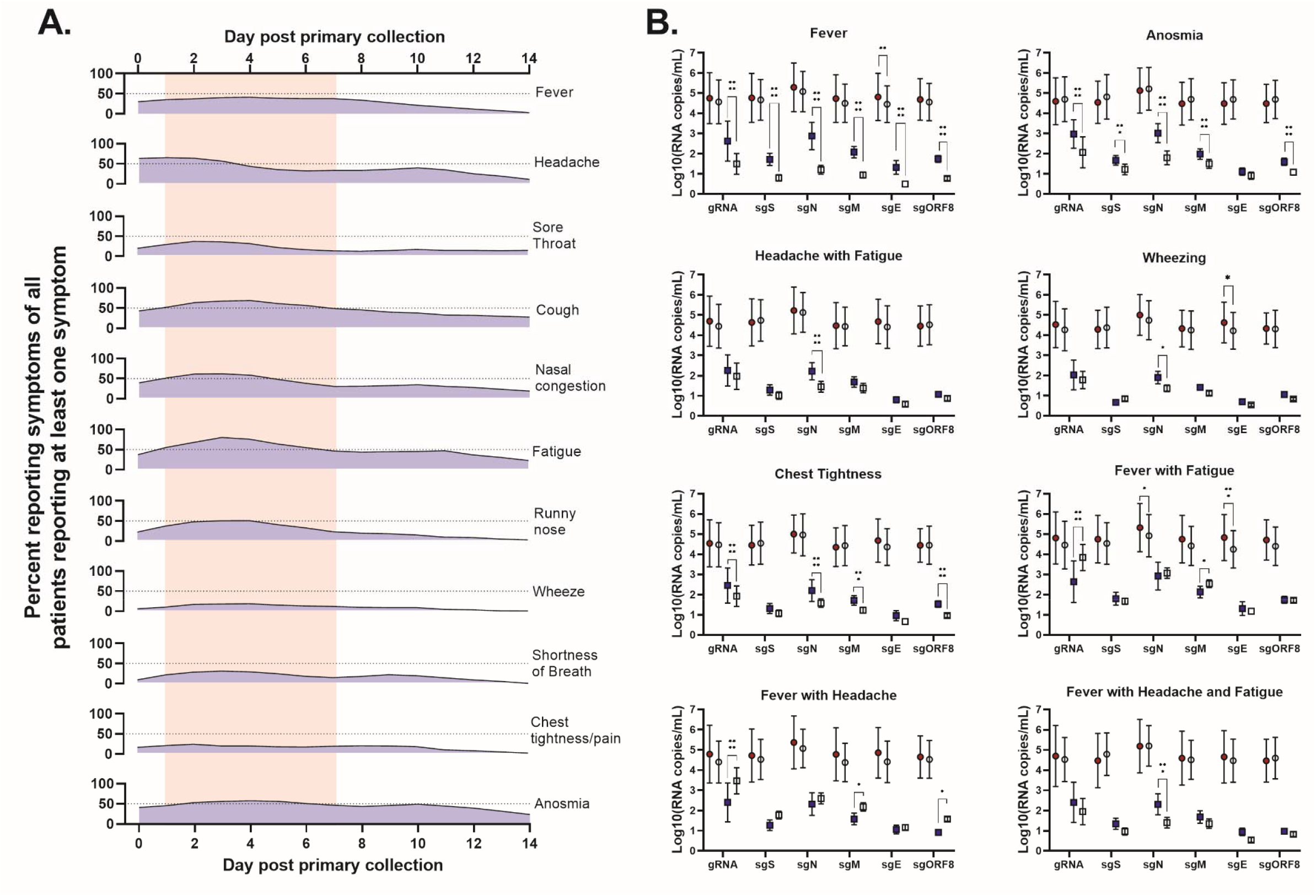
Proportion of patients reporting individual symptoms over time and the relationship between viral RNA levels and symptom presence with and without culturable virus. A) X-axis follows same designation as Figure 3A. The percent of patients reporting a given symptom out of only those who reported one or more symptoms are graphed. Light gray dotted horizontal lines indicate 50%. Solid black lines are 2^nd^ order smoothing (4 neighbors). Transparent red box indicates day of self-collection that patient specimens rendered culturable virus. B) The number of RNA molecules for each analyte were calculated (as described in Figure 2) and grouped by culture result and the presence of symptom(s) indicated at the top of each graph. The weighted average and standard deviation for each data point are shown. All culture positive samples were significantly greater than (****) culture negative specimens, regardless of symptom presence. * = P<0.05; *** = P<0.001; **** = P<0.0001. The same data set used for Figure 3 was used for these analyses.

**Supplemental Figure 4.**
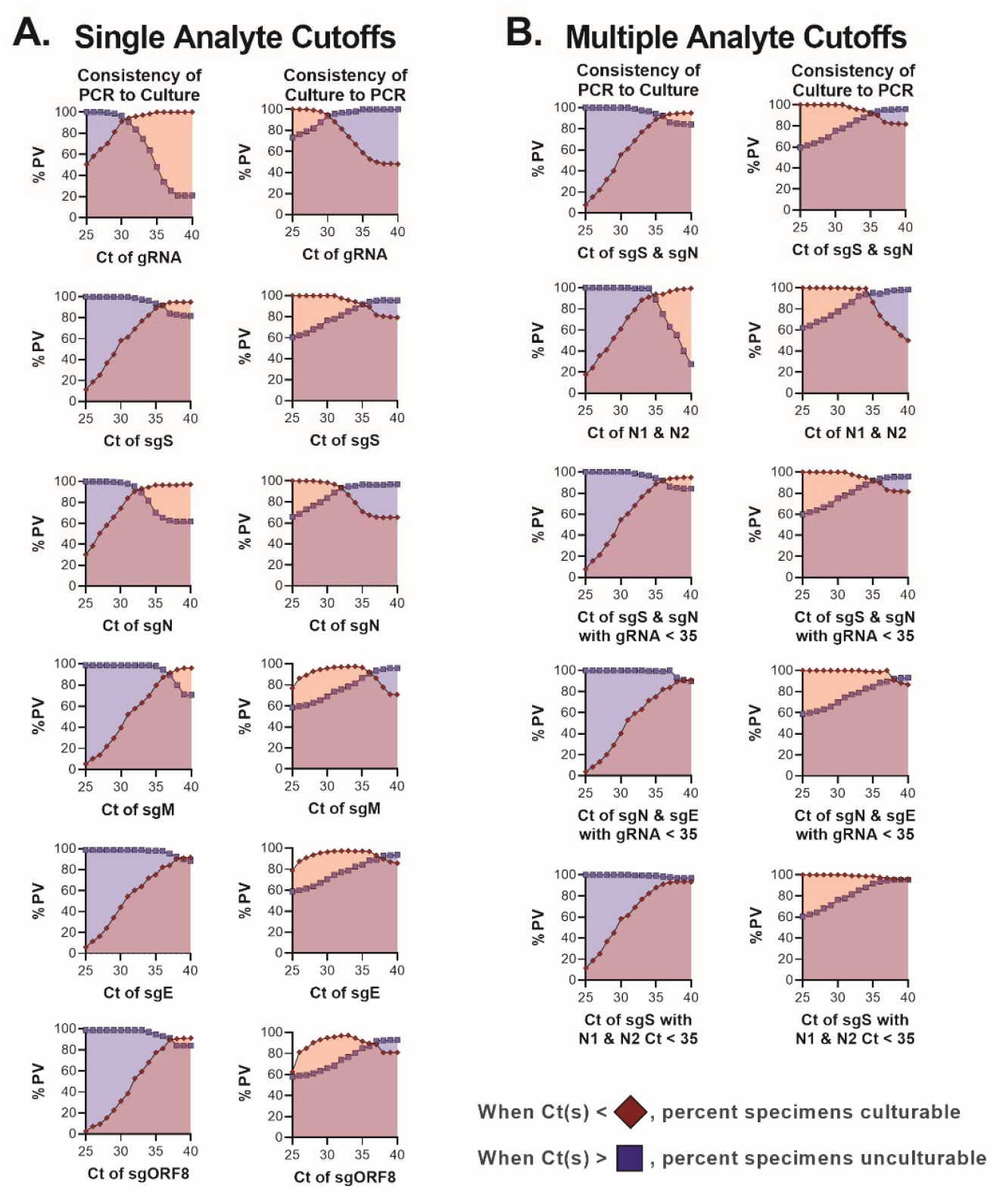
Consistency of single or multiple RNA analytes and culture data. A) To determine the positive and negative predictive values (PPV and NPV, respectively) for the consistency of PCR to culture, the total number of specimens with a Ct value less than or greater than the Ct along the x-axes were divided by the total number of specimens cultured or uncultured, respectively. To determine PPV for the consistency of culture data to PCR, culturable specimens with a Ct less than that along the x-axes were divided by the total number of specimens less than the same Ct cutoff. The NPV for the consistency of culture to PCR was determined by dividing the number of unculturable specimens with a Ct greater than that along the x-axes by the total number of specimens greater than the same Ct cutoff. B) The PPV and NPV using more than one PCR target for consistency between PCR and culture data follow the same equations as in A).

**Supplemental Figure 5:**
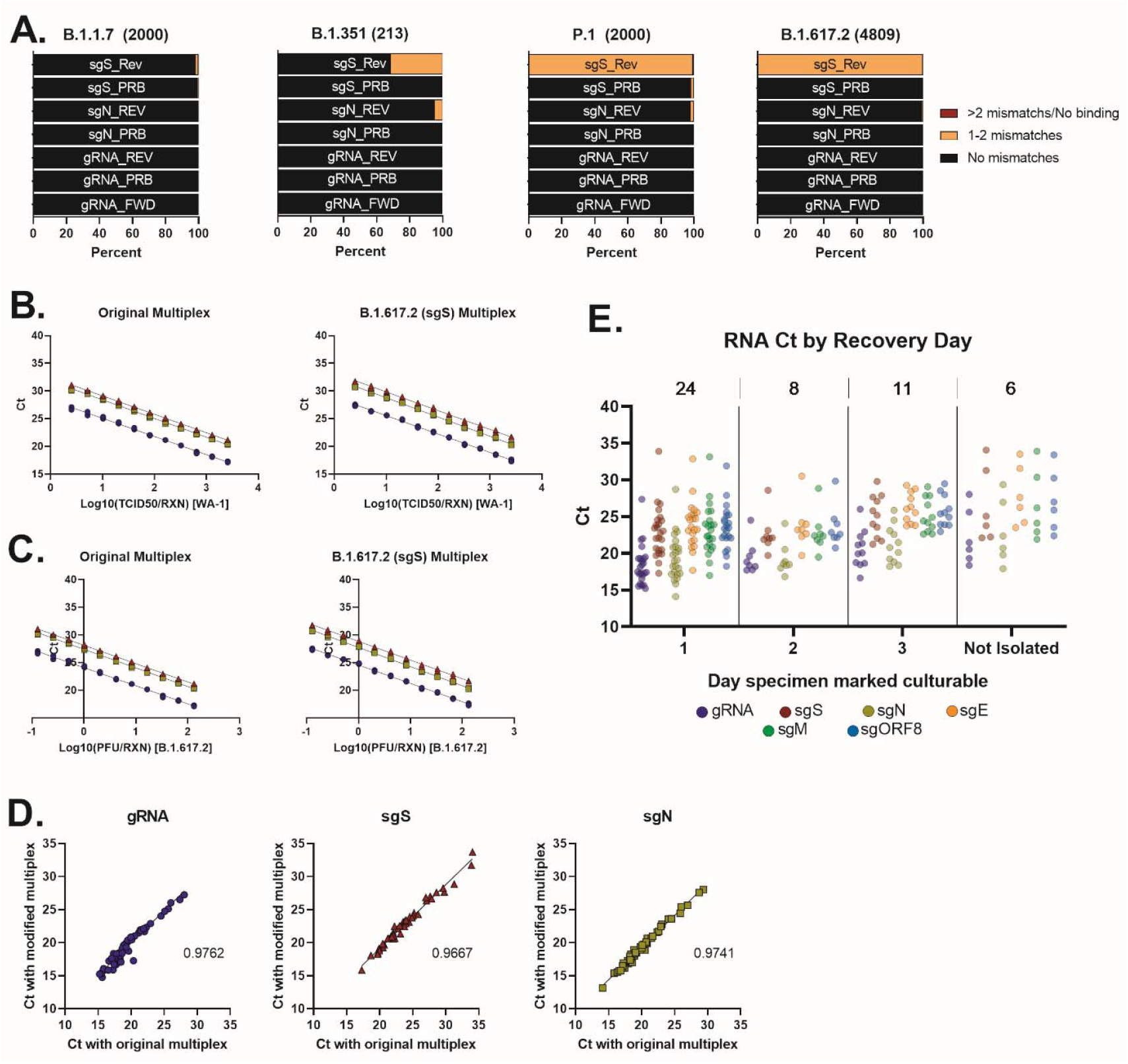
A) Percent of sequences with mismatch in the primer or probe binding region for a given analyte and variant. Listed in parentheses of each graph title are the number of sequences analyzed. B) Standard curves of original multiplex (left) and modified multiplex (right) using serially diluted RNA extracted from supernatant of VeroE6 cells infected with SARS-CoV-2/WA1. C) Standard curves of original multiplex (left) and modified multiplex (right) using serially diluted RNA extracted from supernatant of VeroE6 cells infected with SARS-CoV-2/B.1.617.2. Graphs represent two reactions of each multiplex. D) Original and modified (B.1.617.2) qRT-PCR multiplex mastermixes were assayed against RNA extracted from 50 clinical specimens from a recent infection cluster. The slope of the best-fit line is listed for each graph. E) Ct values for qPCR analytes targeting genomic gRNA and sgRNA are plotted by the indicated group along the x-axis. Above each dot plot are the total number of specimens recovered on a given day post inoculation or not isolated.

**Supplemental Figure 6.**
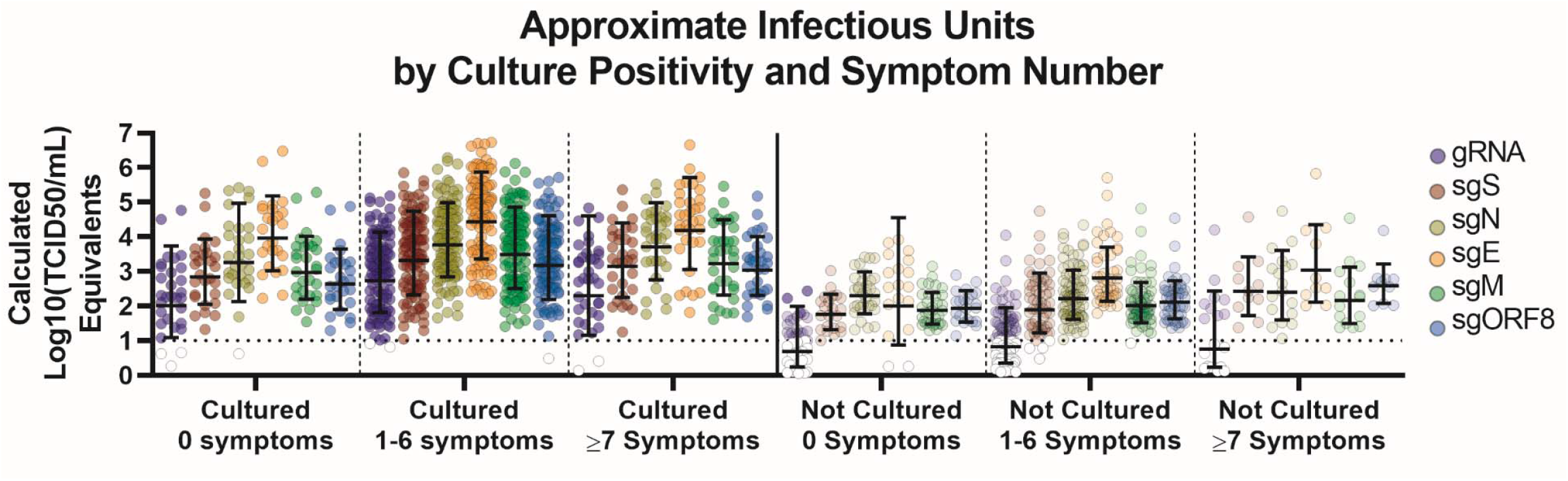
Calculated infectious units by number of symptoms and presence of culturable virus. Standard curves generated using RNA extracted from infected cell culture supernatant for each analyte were used to determine the estimated TCID50 per reaction. Specimens with no detectable RNA are not plotted and did not factor into the mean or 95% confidence interval. Ct values leading to TCID50/mL below the limit of detection (10^1^; dashed line) are white.

