## Supplementary Information for "Surveillance and correlation of SARS-CoV-2 viral RNA, antigen, virus isolation, and self-reported symptoms in a longitudinal study with daily sampling"

**Materials and methods**

***Initial specimen testing and RNA quality assessment, RNA extraction at CDC, and qRT-PCR setup***

Self-collected swabs were stored in traditional viral transport medium and refrigerated until retrieval by study personnel (< five days after collection). Specimens were transported to a central lab at Vanderbilt University Medical Center (Nashville, TN), aliquoted and stored at -80°C. One aliquot was not frozen and used to extract viral RNA with the MagNA Pure LC Total Nucleic Acid Isolation Kit and MagNA PURE LC 2.0 automated extraction platform (Roche). Nucleic acid extracts were tested for SARS-CoV-2 using the 2019-nCoV Assay and associated EUA protocol on StepOnePlus, QuantStudio 3, and QuantStudio 6-Flex real-time PCR systems (ABI). RNA quality assessment for each specimen was based on Ct values generated from the 2019-nCoV Assay. Specimens with cycle threshold (Ct) values ≥ 40 or undetermined for RNaseP were re-extracted and retested, while specimens with RNaseP Ct values < 40 were classified as valid. Specimens producing positive results for the N1 and N2 nucleocapsid gene targets were deemed valid, positive, and final. When N1 or N2 was detected in the absence of the other, qRT-PCR was repeated for both targets. A limited number of specimens positive for N1 or N2, but not both targets, were selected for downstream assay analyses.

Automated extraction of viral RNA from 100 µL respiratory specimens in 400 µL AVL was performed using the QIAamp 96 Virus QIAcube HT kit and system according to manufacturer’s recommendations (Qiagen). RNA extracts were eluted in 100 µL elution buffer into provided elution tubes. RNA extracts not used immediately were stored at -80°C. A mastermix was assembled to include 5 µL TaqPath 1-Step RT-PCR Master Mix (No ROX, ABI), 8 µL H_2_O, and 2 µL of previously assembled primer/probe mixes detailed in Supplementary Table 2. The mastermix was distributed into each well of a 96-well plate prior to adding 5 µL of sample RNA. The reaction plate was sealed with an adhesive optical cover, vortexed briefly, and gently centrifuged to collect well contents. Reaction plates were placed inside of a QuantStudio 6 Pro, equilibrated to 25°C, and incubated at 50°C for 15 minutes. After inactivation of the reverse transcriptase, 45 cycles of 95°C for five seconds followed by 55°C for 30 seconds were carried out. A positive control and no template control were run on every reaction plate. The positive control consisted of RNA extracted from SARS-CoV-2-infected VeroE6 cells spiked with RNA extracted from A549 cells.

*Analysis of disease progression*

Data from each participant were organized longitudinally to analyze RNA levels and symptom data over the time course of infection (Figure 3A). For all study participants with two or more self-collected specimens, the earliest Cx^+^ specimen was arbitrarily assigned one. Subsequent Cx^+^ specimens were given a value based on the day after the first specimen from that study participant was cultured. Unculturable specimens were only considered in this analysis when virus was cultured from an earlier specimen for a given participant and follow the same numerical assignment as Cx^+^ specimens.

*ELISA*

Lyophilized standards were reconstituted, serially diluted, and tested in parallel with specimens from participants. After incubation at 37°C for two hours, plates were treated with a biotin conjugate antibody for one hour followed by streptavidin-HRP for 30 minutes. Reactions were developed by the addition of TMB substrate and absorbance values at 450 and 560 nm were taken with a GloMax Discover instrument (Promega).

**Supplementary Table 1. Primers and probes used for qRT-PCR assay**

| **Assay Identification** | **Primers/probes** | **5’ 🡪 3’** | **Binding position in SARS-CoV-2ª** |
| --- | --- | --- | --- |
| **Leader #1** | Forward | CCAACCAACTTTCGATCTCTTG | 32-53 |
| **Leader #2** | Forward | ACCAACCAACTTTCGATCTCTT | 31-52 |
| **Leader #3** | Forward | AGGTAACAAACCAACCAACTTTC | 22-44 |
| **Leader #4** | Forward | ACCTTCCCAGGTAACAAACC | 14-33 |
| **sg.Spike-1** | Forward | Leader #4 | 14-33 |
|  | Probe | ACCAACTTTCGATCTCTTGTAGATCTGTTC | 35-64 |
|  | Reverse | CTGACTAGAGACTAGTGGCAATAAA | 21,604-21,580 |
| **sg.Spike-2** | Forward | Leader #1 | 32-53 |
|  | Probe | TGCCACTAGTCTCTAGTCAGTGTGT | 21,585-21,609 |
|  | Reverse | GGGTAATTGAGTTCTGGTTGTAAG | 21,637-21,614 |
| **sg.Spike-4** | Forward | CCAGGTAACAAACCAACCAAC | 20-40 |
|  | Probe | CGATCTCCTGTAGATCTGTTCTCTAAACGA | 44-73 |
|  | Reverse | GTAAGATTAACACACTGACTAGAGACTA | 21,618-21,591 |
| **sg.N-1** | Forward | Leader #2 | 31-52 |
|  | Probe | ACGTTTGGTGGACCCTCAGATTCA | 28,319-28,342 |
|  | Reverse | GCGTTCTCCATTCTGGTTACT | 28,369-28,349 |
| **sg.N-2** | Forward | Leader #4 | 14-33 |
|  | Probe | ACCAACTTTCGATCTCTTGTAGATCTGTTC | 35-64 |
|  | Reverse | GAGGGTCCACCAAACGTAAT | 28,335-28,316 |
| **sg.N-4** | Forward | Leader #3 | 22-44 |
|  | Probe | CGCATTACGTTTGGTGGACCCTCA | 28,313-28,336 |
|  | Reverse | TCTGGTTACTGCCAGTTGAAT | 28,358-28,338 |
| **orf1ab-1** | Forward | GAGGCTGTGTGTTCTCTTATGT | 1,485-1,506 |
|  | Probe | AAGTGTGCCTATTGGGTTCCACGT | 1,520-1,543 |
|  | Reverse | CCTTCTCCAACAACACCTGTAT | 1,590-1,569 |
| **orf1ab-2** | Forward | AGAAGTAGGACCTGAGCATAGT | 1,393-1,414 |
|  | Probe | CCATTCTTCGTAAGGGTGGTCGCA | 1,449-1,472 |
|  | Reverse | ACAGCCTCCAAAGGCAATAG | 1,492-1,473 |
| **orf1ab-3** | Forward | CTGAGCATAGTCTTGCCGAATA | 1,404-1,425 |
|  | Probe | ATTCTTCGTAAGGGTGGTCGCACT | 1,451-1,474 |
|  | Reverse | AGAACACACAGCCTCCAAAG | 1,499-1,480 |
| **sg.Envelope-1** | Forward | CAGGTAACAAACCAACC | 21-37 |
|  | Probe | TGTACTCATTCGTTTCGGAAG | 26,246-26,266 |
|  | Reverse | ACTATTAACGTACCTGTCT | 26,285-26,267 |
| **sg.Envelope-3** | Forward | CAAACCAACCAACTTTC | 28-44 |
|  | Probe | ATGTACTCATTCGTTTCGGAAG | 26,245-26,266 |
|  | Reverse | AACTATTAACGTACCTGTC | 26,286-26,268 |
| **sg.Envelope-4** | Forward | TGTAGATCTGTTCTCTAAAC | 52-71 |
|  | Probe | CTCTTCCGAAACGAATGAGTAC | 26,247-26,268 |
|  | Reverse | GTAACTAGCAAGAATACCA | 26,333-26,315 |
| **sg.Membrane-1** | Forward | Leader #3 | 22-44 |
|  | Probe | AAAAGCTCCTTGAACAATGGAACC | 26,563-26,586 |
|  | Reverse | ACAAATCCATGTAAGGAATAGGAAAC | 26,621-26,596 |
| **sg.Membrane-2** | Forward | Leader #2 | 31-52 |
|  | Probe | TGGCAGATTCCAACGGTACT | 26,524-26,543 |
|  | Reverse | AGCTTTTTAAGCTCTTCAACGGTA | 26,569-26,546 |
| **sg.Membrane-3** | Forward | Leader #1 | 32-53 |
|  | Probe | TCCAACGGTACTATTACCGTTGAAGA | 26,532-26,557 |
|  | Reverse | GTTCCATTGTTCAAGGAGCTTTT | 26,585-26,563 |
| **sg.orf8-1** | Forward | Leader #2 | 31-52 |
|  | Probe | TGAAATTTCTTGTTTTCTTAGGAATCATCAC | 27,895-27,925 |
|  | Reverse | CTTGGTGAAATGCAGCTACAG | 27,948-27,928 |
| **sg.orf8-2** | Forward | Leader #2 | 31-52 |
|  | Probe | ACAACTGTAGCTGCATTTCACC | 27,923-27,944 |
|  | Reverse | GTTGAGTACATGACTGTAAACTACATTC | 27,975-27,948 |
| **sg.orf8-3** | Forward | Leader #3 | 22-44 |
|  | Probe | CTGCATTTCACCAAGAATGTAGTTTACA | 27,934-27,961 |
|  | Reverse | CTACATATGGTTGATGTTGAGTACATG | 27,990-27,964 |
| **sg.Spike_#2Δ-1^b^** | Forward | Leader #3 | 22-44 |
|  | Probe | TGCCACTAGTCTTTAGTCAGTGTGT | 21,585-21,609 |
|  | Reverse | GGGTAATTGAGTTCTGGTTCTAAG | 21,637-21,614 |
| **sg.Spike_#2Δ-4^b^** | Forward | Leader #3 | 22-44 |
|  | Probe | TGCCACTAGTCTTTAGTCAGTGTGT | 21,585-21,609 |
|  | Reverse | GTGAAAGAATTAGTGTATGCAGG | 21,638-21,660 |
| **sg.Spike_#2Δ-5^b^** | Forward | Leader #3 | 22-44 |
|  | Probe | TGCCACTAGTCTTTAGTCAGTGTGT | 21,585-21,609 |
|  | Reverse | TGTGAAAGAATTAGTGTATGCAG | 21,660-21,638 |
| ª Binding location of primers when aligned to SARS-CoV-2/Wuhan-Hu-1 (NCBI Reference Sequence: NC_045512.2)  **^b^** Binding location of primers when aligned to SARS-CoV-2/B.1.617.2 | | | |

**Supplementary Table 2. Frequently used cell culture reagents**

| **Component** | **Manufacturer (Product #)** |
| --- | --- |
| **Dulbecco’s Modified Eagle Medium (DMEM)** | **Gibco (11965-092)** |
| **Modified Eagle Medium (MEM)** | **Gibco (11935-046)** |
| **Fetal Bovine Serum (FBS)** | **Gibco (A3160701)** |
| **Geneticin** | **Gibco (10131027)** |
| **Antibiotic-antimycotic** | **Gibco (15240062)** |
| **Penicillin/Streptomycin (P/S)** | **Gibco (16140148)** |
| **Cellulose** | **Sigma-Aldrich (435244)** |
| **L-Glutamine** | **Gibco (25030164)** |
| **Vero E6-TMPRSS2**  **maintenance media** | **DMEM, 10% FBS,**  **0.5 mg/mL geneticin** |
| **Vero E6-TMPRSS2**  **infection media** | **DMEM, 2% FBS,**  **1% antibiotic-antimycotic** |
| **Virus diluent** | **DMEM, 2% FBS, 1% P/S** |
| **1% Cellulose solution** | **MEM, 5% FBS, 2% P/S, 1% L-glutamine** |

**Supplementary Table 3. Composition of multiplex primer and probe mixes.**

| **Multiplex #1** | | | |
| --- | --- | --- | --- |
| **Assay Identification** | **Primers/probes*** | **^ї^ 5’ 🡪 3’** | **Binding position in SARS-CoV-2ª** |
| Leader-3 | Forward (750) | AGGTAACAAACCAACCAACTTTC | 22-44 |
| sg.N-1 | Reverse (400) | GCGTTCTCCATTCTGGTTACT | 28,369-28,349 |
|  | Probe (125) | (Cy5) ACGTTTGGTGGACCCTCAGATTCA | 28,319-28,342 |
| sg.Spike-2 | Reverse (400) | GGGTAATTGAGTTCTGGTTGTAAG | 21,637-21,614 |
|  | Probe (125) | (FAM) TGCCACTAGTCTCTAGTCAGTGTGT | 21,585-21,609 |
| orf1ab-1 | Forward (500) | GAGGCTGTGTGTTCTCTTATGT | 1,485-1,506 |
|  | Probe (125) | (SUN) AAGTGTGCCTATTGGGTTCCACGT | 1,520-1,543 |
|  | Reverse (500) | CCTTCTCCAACAACACCTGTAT | 1,590-1,569 |
| RNaseP | Forward (500) | AGATTTGGACCTGCGAGCG |  |
|  | Probe (125) | (TAMN) TTCTGACCTGAAGGCTCTGCGCG |  |
|  | Reverse (500) | GAGCGGCTGTCTCCACAAGT |  |
| * Quantity of primer or probe in nm listed in parentheses.  ^ї^ Probes labeled with the indicated molecule in parentheses. sg.Spike-2 and orf1ab probes were internally quenched with ZEN, sg.N-1 with TAO, and no quencher was used for RNaseP. RNaseP and sg.N-1 were labeled at the 3’ end with IAbRQSp while sg.Spike-2 and orf1ab-1 were labeled with IABkFQ at the 3’ end. Integrated DNA Technologies Inc. (IDT) <http://www.idtdna.com> (or equivalent) – RNase-Free and HPLC Purified  ª Binding location of primers when aligned to SARS-CoV-2/Wuhan-Hu-1 (NCBI Reference Sequence: NC_045512.2) | | | |

| **Multiplex #2** | | | |
| --- | --- | --- | --- |
| **Assay Identification** | **Primers/probes*** | **^ї^ 5’ 🡪 3’** | **Binding position in SARS-CoV-2ª** |
| Leader-1 | Forward (750) | CCAACCAACTTTCGATCTCTTG | 32-53 |
| sg.Envelope-4 | Reverse (500) | GTAACTAGCAAGAATACCA | 26,315-26,333 |
|  | Probe (125) | (FAM) CTCTTCCGAAACGAATGAGTAC | 26,247-26,268 |
| sg.Membrane-2 | Reverse (500) | AGCTTTTTAAGCTCTTCAACGGTA | 26,569-26,546 |
|  | Probe (125) | (CY5) TGGCAGATTCCAACGGTACT | 26,524-26,543 |
| sg.orf8-2 | Reverse (500) | GTTGAGTACATGACTGTAAACTACATTC | 27,975-27,948 |
|  | Probe (125) | (SUN) ACAACTGTAGCTGCATTTCACC | 27,924-27,945 |
| * Quantity of primer or probe in nm listed in parentheses.  ^ї^ Probes labeled with the indicated molecule in parentheses Integrated DNA Technologies Inc. (IDT) <http://www.idtdna.com> (or equivalent) – RNase-Free and HPLC Purified  ª Binding location of primers when aligned to SARS-CoV-2/Wuhan-Hu-1 (NCBI Reference Sequence: NC_045512.2) | | | |

**Supplementary Table 4: Calculated RNA copies between culturable and unculturable specimens that are antigen positive or negative.**

|  | | **gRNA** | **sgS** | **sgN** | **sgE** | **sgM** | **sgORF8** |
| --- | --- | --- | --- | --- | --- | --- | --- |
| **Cx^+^, Ag^+^** | **Log10(RNA copies/mL)** | 4.55 ± 1.12 | 5.04 ± 1.07 | 5.23 ± 1.08 | 5.08 ± 1.13 | 4.68 ± 1.04 | 4.63 ± 1.01 |
|  | **qRT-PCR+** | 168/168 (100%) | 162/168 (96.4%) | 165/168 (98.2%) | 154/168 (91.7%) | 161/168 (95.8%) | 155/168 (92.3%) |
| **Cx^+^, Ag^-^** | **Log10(RNA copies/mL)** | 2.67 ± 0.9 | 5.10 ± 1.22 | 3.57 ± 1.14 | 5.17 ± 1.08 | 3.84 ± 1.16 | 3.69 ± 1.10 |
|  | **qRT-PCR+** | 11/11 (100%) | 8/11 (72.7%) | 9/11 (81.8%) | 9/11 (81.8%) | 10/11 (90.9%) | 8/11 (72.7%) |
| **Significance** | | ** | NS | ** | NS | NS | NS |
| **Cx^-^, Ag^+^** | **Log10(RNA copies/mL)** | 1.68 ± 0.57 | 3.11 ± 0.47 | 3.08 ± 0.32 | 3.05 ± 0.62 | 2.90 ± 0.30 | 2.88 ± 0.14 |
|  | **qRT-PCR+** | 174/194 (89.7%) | 81/194 (41.8%) | 122/194 (62.9%) | 65/194 (33.5%) | 101/194 (52.1%) | 74/194 (38.1%) |
| **Cx^-^, Ag^-^** | **Log10(RNA copies/mL)** | 1.88 ± 0.76 | 3.01 ± 0.28 | 3.26 ± 0.47 | 3.31 ± 0.35 | 2.80 ± 0.29 | 3.11 ± 0.25 |
|  | **qRT-PCR+** | 41/56 (73.2%) | 10/56 (17.9%) | 15/56 (26.8%) | 4/56 (7.1%) | 15/56 (26.8%) | 8/56 (14.3%) |
| **Significance** | | NS | NS | NS | NS | NS | ** |
| Cx: Culture isolation  Ag: Antigen  **: P value <0.0001 when comparing the Ct value of the indicated RNA species for culture result with or without positive antigen detection  NS: Not significant | | | | | | | |

**Supplementary Table 5: Ct values of specimens with unexpected results: high Ct value and culturable versus low Ct value and unculturable**

| **High Ct; isolated** | | | | | | | | | | | | | | | | | | | | | | | | | | | |
| --- | --- | --- | --- | --- | --- | --- | --- | --- | --- | --- | --- | --- | --- | --- | --- | --- | --- | --- | --- | --- | --- | --- | --- | --- | --- | --- | --- |
| **Sample ID** | **CDC-N1** | **CDC-N2** | | **gRNA** | | **sgS** | | **sgN** | **sgE** | | **sgM** | **sgORF8** | | **Reported Symptoms** | | | | | | | | | | | | | **Antigen** |
|  |  |  |  |  |  |  |  |  |  |  |  |  |  | **1** | **2** | **3** | **4** | **5** | **6** | | | **7** | **8** | **9** | **10** | **11** |  |
| **1** | 36.76 | ND | | 33.15 | | ND | | ND | ND | | ND | ND | | X | X | X | X | X | X | | | X | X | X | X | X | No |
| **2** | 39.18 | 38.29 | | 34.05 | | ND | | ND | 37.99 | | 37.82 | ND | | X | X |  | X | X | X | | | X | X | X | X | X | No |
| **3** | 34.94 | 36.37 | | 30.97 | | ND | | 34.88 | ND | | 37.90 | ND | | X |  |  |  | X | X | | |  |  |  | X |  | No |
| **4** | 34.35 | 36.16 | | 33.63 | | 34.75 | | 30.04 | 35.98 | | 32.42 | 34.21 | |  |  |  |  |  |  | | |  |  |  |  |  | Yes |
| **5** | 35.55 | 36.13 | | 32.45 | | ND | | 34.77 | ND | | 38.04 | ND | |  |  |  |  |  |  | | |  |  |  |  |  | Yes |
| **6** | 35.37 | 37.32 | | 34.97 | | ND | | ND | ND | | ND | ND | |  |  |  |  |  |  | | |  |  |  |  |  | Yes |
| **7** | 34.57 | 36.23 | | 30.62 | | 36.65 | | 33.99 | 38.00 | | 36.95 | 36.01 | | X | X |  |  | X | X | | | X | X |  |  | X | Yes |
| **8** | 34.11 | 37.21 | | 29.88 | | 34.22 | | 32.43 | ND | | ND | ND | | X |  |  |  |  | X | | |  |  |  |  |  | Yes |
| **9** | 34.41 | 38.28 | | 31.79 | | ND | | 34.63 | ND | | 38.52 | ND | | X | X | X | X | X | X | | |  |  |  |  | X | Yes |
| **10** | 35.94 | 37.49 | | 34.84 | | ND | | ND | ND | | ND | ND | |  |  | X |  | X |  | | |  |  |  |  |  | Yes |
| **Low Ct; not isolated** | | | | | | | | | | | | | | | | | | | | | | | | | | | |
| **Sample ID** | **CDC-N1** | **CDC-N2** | | **gRNA** | | **sgS** | | **sgN** | **sgE** | | **sgM** | **sgORF8** | | **Reported Symptoms** | | | | | | | | | | | | | **Antigen** |
|  |  |  |  |  |  |  |  |  |  |  |  |  |  | **1** | **2** | **3** | **4** | **5** | **6** | | | **7** | **8** | **9** | **10** | **11** |  |
| **1** | 27.80 | 28.42 | | 25.87 | | 32.51 | | 30.25 | 33.88 | | 34.41 | 35.37 | |  |  |  | X |  |  | | |  |  |  |  |  | Yes |
| **2** | 27.90 | 28.32 | | 26.31 | | 30.55 | | 30.46 | 33.27 | | 34.05 | 35.66 | |  | X | X | X | X | X | | |  |  |  |  |  | Yes |
| **3** | 29.81 | 31.13 | | 26.68 | | 32.53 | | 30.86 | 35.63 | | 34.60 | 34.28 | |  |  |  |  |  |  | | |  |  |  |  |  | Yes |
| **4** | 29.71 | 30.90 | | 25.92 | | 30.89 | | 28.87 | 33.00 | | 32.42 | 32.34 | |  |  |  |  |  |  | | |  |  |  |  |  | Yes |
| **5** | 29.47 | 29.90 | | 25.69 | | 31.36 | | 29.43 | 33.02 | | 33.03 | 32.69 | |  |  |  |  | X | X | | |  |  |  | X |  | Yes |
| **6** | 28.56 | 31.68 | | 26.79 | | 31.10 | | 27.61 | 32.32 | | 32.56 | 31.81 | | X | X |  |  | X | X | | |  |  |  | X |  | Yes |
| **7** | 29.64 | 27.90 | | 26.10 | | 30.49 | | 28.55 | 32.11 | | 35.18 | 34.72 | | X |  |  |  |  | X | | |  |  |  |  |  | Yes |
| ^1^Fever, ^2^Cough, ^3^Sore throat, ^4^Runny nose, ^5^Nasal congestion, ^6^Fatigue, ^7^Wheeze,^8^Short breath, ^9^Chest pain/tightness, ^10^Loss of taste/smell, ^11^Headache | | | | | | | | | | | | | | | | | | | | |  | | | | | | |

**Supplementary Table 6. Characteristics of index participants by number of days culture positive**

|  |  | **Number of days from illness onset to last culture positive specimen collected** | | |  |  |
| --- | --- | --- | --- | --- | --- | --- |
|  | **Total**  **N=39** | **Never**  **N =7** | **< 6 days**  **N=16** | **≥ 6 days**  **N=16** | **p^1^** | **p^2^** |
| ***Index Case Characteristics*** | | | | | | |
|  | N (%) | N (%) | N (%) | N (%) |  |  |
| **Demographics** | | | | | | |
| **Sex** | | | | | 0.0307 | 0.0944 |
| **Male** | 15 (38.46) | 0 (0.00) | 6 (37.50) | 9 (56.25) |  |  |
| **Female** | 24 (61.54) | 7 (100.00) | 10 (62.50) | 7 (43.75) |  |  |
| **Age** | | | | | 0.2442 | 0.4632 |
| **<11** | 0 (0.00) | 0 (0.00) | 0 (0.00) | 0 (0.00) |  |  |
| **12-17** | 1 (2.56) | 1 (14.29) | 0 (0.00) | 0 (0.00) |  |  |
| **18-30** | 17 (43.59) | 4 (57.14) | 8 (50.00) | 5 (31.25) |  |  |
| **30-50** | 13 (33.33) | 2 (28.57) | 4 (25.00) | 7 (43.75) |  |  |
| **50-65** | 7 (17.95) | 0 (0.00) | 3 (18.75) | 4 (25.00) |  |  |
| **65+** | 1 (2.56) | 0 (0.00) | 1 (6.25) | 0 (0.00) |  |  |
| **Race** | | | | | | |
| **White** | 31 | 4 | 14 | 13 | 0.1368 | 1.0000 |
| **Black** | 2 | 1 | 0 | 1 | 0.3306 | 1.0000 |
| **Asian** | 1 | 0 | 1 | 0 | 1.0000 | 1.0000 |
| **Native Hawaiian/Pacific Islander** | 0 | 0 | 0 | 0 | N/A | N/A |
| **American Indian/ Alaskan Native** | 0 | 0 | 0 | 0 | N/A | N/A |
| **Other** | 6 | 2 | 2 | 2 | 0.2902 | 1.0000 |
| **Ethnicity** | | | | | 1.0000 | 1.0000 |
| **Non-Hispanic** | 28 (71.79) | 5 (71.43) | 11 (68.75) | 12 (75.00) |  |  |
| **Hispanic** | 11 (28.21) | 2 (28.57) | 5 (31.25) | 4 (25.00) |  |  |
| **Mitigation Factors** | | | | | | |
| **Mask Use** | | | | | 0.5986 | 0.9592 |
| **Never** | 12 (30.77) | 2 (28.57) | 6 (37.50) | 4 (25.00) |  |  |
| **Some of the time** | 17 (43.59) | 3 (42.86) | 6 (37.50) | 8 (50.00) |  |  |
| **Most of the time** | 5 (12.82) | 2 (28.57) | 1 (6.25) | 2 (12.50) |  |  |
| **Always** | 5 (12.82) | 0 (0.00) | 3 (18.75) | 2 (12.50) |  |  |
| **Separate Room** | | | | | 0.7333 | 0.6664 |
| **Never** | 6 (15.38) | 2 (28.57) | 2 (12.50) | 2 (12.50) |  |  |
| **Some of the time** | 6 (15.38) | 1 (14.29) | 3 (18.75) | 2 (12.50) |  |  |
| **Most of the time** | 4 (10.26) | 0 (0.00) | 1 (6.25) | 3 (18.75) |  |  |
| **Always** | 23 (58.97) | 4 (57.14) | 10 (62.50) | 9 (56.25) |  |  |
| **Separate Bathroom** | | | | | 0.0627 | 0.6078 |
| **Never** | 10 (25.64) | 2 (28.57) | 4 (25.00) | 4 (25.00) |  |  |
| **Some of the time** | 5 (12.82) | 1 (14.29) | 2 (12.50) | 2 (12.50) |  |  |
| **Most of the time** | 6 (15.38) | 3 (42.86) | 2 (12.50) | 1 (6.25) |  |  |
| **Always** | 18 (46.15) | 1 (14.29) | 8 (50.00) | 9 (56.25) |  |  |
| **Eat Separate** | | | | | 0.0520 | 0.6304 |
| **Never** | 4 (10.26) | 1 (14.29) | 2 (12.50) | 1 (6.25) |  |  |
| **Some of the time** | 12 (30.77) | 3 (42.86) | 4 (25.00) | 5 (31.25) |  |  |
| **Most of the time** | 8 (20.51) | 3 (42.86) | 3 (18.75) | 2 (12.50) |  |  |
| **Always** | 15 (38.46) | 0 (0.00) | 7 (43.75) | 8 (50.00) |  |  |
| **Hand Washing/Hand Sanitizer** | | | | | 0.1089 | 0.4869 |
| **Never** | 2 (5.13) | 0 (0.00) | 0 (0.00) | 2 (12.50) |  |  |
| **Some of the time** | 4 (10.26) | 1 (14.29) | 2 (12.50) | 1 (6.25) |  |  |
| **Most of the time** | 6 (15.38) | 3 (42.86) | 1 (6.25) | 2 (12.50) |  |  |
| **Always** | 27 (69.23) | 3 (42.86) | 13 (81.25) | 11 (68.75) |  |  |
| **Preexisting Medical Conditions** | | | | | | |
| **Any Preexisting Condition** | 10 (25.64) | 2 (28.57) | 5 (31.25) | 3 (18.75) | 1.0000 | 0.4798 |
| **Symptoms During the Illness Episode** | | | | | | |
| **Fever** | 33 (84.62) | 6 (85.71) | 15 (93.75) | 12 (75.00) | 1.0000 | 0.2053 |
| **Cough** | 32 (82.05) | 5 (71.43) | 13 (81.25) | 14 (87.50) | 0.5876 | 0.6776 |
| **Sore Throat** | 21 (53.85) | 2 (28.57) | 10 (62.50) | 9 (56.25) | 0.2155 | 1.0000 |
| **Runny Nose** | 27 (69.23) | 4 (57.14) | 11 (68.75) | 12 (75.00) | 0.6536 | 0.7262 |
| **Nasal Congestion** | 31 (79.49) | 5 (71.43) | 14 (87.50) | 12 (75.00) | 0.6170 | 0.6937 |
| **Fatigue** | 36 (92.31) | 6 (85.71) | 15 (93.75) | 15 (93.75) | 0.4573 | 1.0000 |
| **Wheezing** | 15 (38.46) | 3 (42.86) | 7 (43.75) | 5 (31.25) | 1.0000 | 0.5166 |
| **Shortness of Breath** | 23 (58.97) | 6 (85.71) | 8 (50.00) | 9 (56.25) | 0.2055 | 1.0000 |
| **Loss of taste/smell** | 29 (78.38) | 7 (100.00) | 13 (81.25) | 9 (64.29) | 0.3079 | 0.2152 |
| **Chest Tightness/Pain** | 21 (53.85) | 5 (71.43) | 9 (56.25) | 7 (43.75) | 0.4179 | 0.3422 |
| **Headache** | 35 (92.11) | 7 (100.00) | 16 (100.00) | 12 (80.00) | 1.0000 | 0.0539 |
| **Transmission** | | | | | | |
| **Transmitted to at least one household contact** | 25 (64.10) | 5 (71.43) | 10 (62.50) | 10 (62.50) | 1.0000 | 1.0000 |
| ^1^p-value for fisher’s exact test between never culture positive and at least one day culture positive  ^2^p-value for fisher’s exact test between < 6 days culture positive (including never) and ≥ 6 days culture positive | | | | | | |
